# Replicating a COVID-19 study in a national England database to assess the generalisability of research with regional electronic health record data

**DOI:** 10.1101/2024.08.06.24311538

**Authors:** Richard Williams, David Jenkins, Thomas Bolton, Adrian Heald, Mehrdad A Mizani, Matthew Sperrin, Niels Peek, the CVD-COVID-UK/COVID-IMPACT Consortium

## Abstract

**Objectives:** To assess the degree to which we can replicate a study between a regional and a national database of electronic health record data in the United Kingdom.

**Design:** A replication of a retrospective cohort study.

**Setting:** Observational EHR data from primary and secondary care sources in the UK. The original study used data from a large, urbanised region (Greater Manchester Care Record, Greater Manchester, UK). This replication study used a national database covering the whole of England, UK (NHS England’s Secure Data Environment service for England, accessed via the BHF Data Science Centre’s CVD-COVID-UK/COVID-IMPACT Consortium).

**Participants:** Individuals with a diagnosis of T1D or T2D prior to a positive COVID-19 test result. The matched controls (3:1) were individuals who had a positive COVID-19 test result, but who did not have a diagnosis of diabetes on the date of their positive COVID-19 test result. Matching was done on age at COVID-19 diagnosis, sex and approximate date of COVID-19 test.

**Primary and secondary outcome measures:** Hospitalization within 28 days of a positive COVID-19 test.

**Results:** We found that many of the effect sizes did not show a statistically significant difference. Where effect sizes were statistically significant in the regional study, then they remained significant in the national study and the effect size was the same direction and of similar magnitude.

**Conclusions:** There is some evidence that the findings from studies in smaller regional datasets can be extrapolated to a larger, national setting. However, there were some significant differences and therefore replication studies remain an essential part of healthcare research.

**Strengths and limitations of this study:**

- The same team performed the original study and this replication study
- The underlying data sources, while similar, had differences that may have affected the results
- The focus of replication was a single outcome for a single condition and may not generalise to other disease areas

## 1 Introduction

Observational studies using electronic health record (EHR) data are a critical component of the evidence base in population health and epidemiology. However, their findings carry less weight in evidence-based medicine when compared with more conclusive results such as those from randomised control trials. This is partly due to concerns about generalizability and the potential for confounding biases. Replication, the process of repeating a study with a different population or data source, is crucial for strengthening the evidence base in observational research. Successful replication of findings can significantly improve our confidence in their validity and generalizability, leading to a more robust foundation for policy and clinical practice decisions.

Reproducibility is one of the greatest challenges in the area of observational studies [1,2]. Goodman et al. define three terms for discussing research reproducibility: methods reproducibility, results reproducibility and inferential reproducibility [3]. Methods reproducibility is the degree to which a publication includes sufficient information such that other researchers could repeat the analysis. Results reproducibility is the degree to which other researchers can achieve the same results.

We have previously published a study where we compared hospitalization rates of patients in Greater Manchester (GM) with type 1 diabetes (T1D) or type 2 diabetes (T2D) after contracting COVID-19 when compared with age and sex matched controls [4]. The study reported that following confirmed infection with COVID-19, a number of factors are associated with increased levels of hospitalization in individuals with T1D and T2D. For patients with T1D, older age, increased social disadvantage, and having hypertension or COPD were associated with an increased risk of hospitalization. Other factors were non-significant, potentially due to the small population size. Patients with T2D had the same risk factors as patients with T1D, but with the addition that male sex, non-white ethnicity and severe mental illness had an increased risk of hospitalization, while taking metformin and low cholesterol levels were associated with a reduced risk of hospitalization. In this study we will attempt to replicate these findings in a national database covering the whole of England.

For this replication study, methods reproducibility should have been trivial as we performed the original analysis. However, this was not the case and in a separate paper we discuss the methodological factors that make replication problematic, such as differences in the governance, the data structure and the data processing [5]. Inferential reproducibility is not possible as it is the degree to which different researchers reach the same conclusions from similar results. Therefore, in this paper our objective is to assess the degree to which we can achieve results reproducibility between a regional and a national database of electronic health record data in the United Kingdom (UK).

If results reproducibility can be achieved then this will provide evidence that, under certain circumstances, scientific conclusions drawn from regional datasets can be extrapolated nationally.

## 2 Methods

### 2.1 Study design

This is a replication of a retrospective cohort study using observational EHR data from primary and secondary care sources in the UK.

### 2.2 Data sources

The data for the original study were from the Greater Manchester Care Record (GMCR). The GMCR is a shared care record containing primary and secondary care data for the residents of Greater Manchester. The database contains all primary care data, and all hospital admission data, for patients registered to a GP in GM who have not opted out of data sharing.

The data for this replication study were from the NHS England National Secure Data Environment (National SDE). The National SDE provides access to a range of national data sets relating to healthcare. Data were made available for COVID-19 research through the CVD-COVID-UK/COVID-IMPACT Consortium which is coordinated by the BHF Data Science Centre and led by Health Data Research UK. The data used for this study were: primary care data from the General Practice Extraction Service (GPES) Data for Pandemic Planning and Research (GDPPR) [6]; secondary care data from Hospital Episode Statistics (HES) Admitted Patient Care (APC) [7]; and COVID-19 test data from the Second Generation Surveillance System (SGSS) data set [8].

### 2.3 Setting

In the original study, all patients from Greater Manchester (population 2.8m) with a positive COVID-19 test in their primary care record between 1^st^ January 2020 (month of first UK cases of COVID-19) and 31^st^ May 2021 were eligible.

In this replication study we have a larger data source. Patients are now from the whole of England (population 54m after excluding ∼1.3m opt-outs) [9]. COVID-19 tests are from the SGSS, in addition to those from the primary care record. The date range is now 1^st^ January 2020 to 1^st^ January 2023. The SGSS contains all community COVID-19 test results and so is more complete than the COVID-19 results that appear in a patient’s primary care record.

### 2.4 Approach

We conducted two analyses. Our initial GM study relied on COVID-19 test results that appeared in the primary care record. Therefore, the first analysis was an attempt to reproduce the results of our original study, by only using the COVID-19 test data from the primary care part of the National SDE.

The second analysis used the COVID-19 test data from the SGSS, in addition to the primary care data, as this is how researchers using the national SDE would obtain COVID-19 test results.

### 2.5 Study population

For all analyses the main cohort was defined as patients with a diagnosis of T1D or T2D prior to a positive COVID-19 test result. The controls were patients who had a positive COVID-19 test result, but who did not have a diagnosis of diabetes on the date of their positive COVID-19 test result. Each patient in the main cohort was matched with up to 3 controls. Controls were not reused for multiple patients. Matching was done on age at COVID-19 diagnosis, sex and approximate date (within 2 weeks either side) of COVID-19 test. The date of COVID-19 test is important as outcomes differ depending on the particular wave or variant of COVID-19 that they contracted.

### 2.6 Variables

The outcome is all-cause hospitalisation within 28 days of a positive COVID-19 test. The original study used feeds of admissions data from each hospital within GM. This replication study used the APC table from HES data.

The covariates are a subset of those from the original study. They are: year of birth; sex; ethnicity; deprivation via the Townsend score; latest values prior to the COVID-19 result for BMI, Hba1c, cholesterol (total, LDL and HDL) and eGFR; smoking status; whether the patient has COPD, asthma, a severe mental illness or hypertension; whether the patient is currently prescribed an ACE inhibitor or ARB, aspirin, clopidogrel or metformin. The covariates in the original study that were not available for this replication study were: latest values prior to COVID-19 result for vitamin D, testosterone and sex hormone binding globulin. These biomarkers were not available in the GDPPR dataset, which contains a subset of SNOMED concepts from a patient’s primary care record, and therefore were excluded from the analysis.

### 2.7 Statistical methods

The original study’s objective was to identify potential factors relating to an increased likelihood of hospital admission in individuals with diabetes, to assess the difference in risk between individuals with and without diabetes and to investigate if any difference in risk could be explained by routinely measured factors. The statistical analysis methods are an exact replication of the previous study [4]. A brief overview is as follows.

Modelling was conducted using conditional logistic regression with hospitalisation within 28 days of a positive COVID-19 test as the outcome. We analysed the individuals with diabetes, without the matched controls, using a univariable logistic regression for each factor in turn, for the two groups (T1D and T2D) separately. We then fitted a multivariable model using the patients with diabetes and their controls, with diabetes diagnosis as a covariate and adjusting for other factors.

Following these analyses, we compared the national effect sizes and odds ratios (ORs) to our previous work from the GMCR dataset. In addition to a descriptive comparison, we also calculated a conservative 95% confidence interval for the difference between the odds ratios to find whether there was a statistically significant difference between the effect sizes between GM and the national data.

This analysis was performed according to a pre-specified analysis plan published on GitHub, along with the phenotyping and analysis code (https://github.com/BHFDSC/CCU040_01).

## 3 Results

Our objective is to demonstrate the degree to which results reproducibility can be achieved. Therefore, all odds ratios and confidence intervals (CIs) are displayed visually and discussed descriptively. Full tables with numeric data are available in the supplementary material (Tables S1-S4).

### 3.1 Population comparison

Both national analyses benefited from a much larger population. The original GM study had 862 patients with T1D and a positive COVID-19 test result, while the first national analysis had 38,523, and the second had 77,392 (Table 1). The original study had 13,225 patients with T2D and a positive COVID-19 test result, while the first national analysis had 448,829, and the second had 836,532 (Table 2). We have previously published a clinical paper focussing on the individuals with T1D [10].

**Table 1.** characteristics of the individuals with type 1 diabetes (T1D) and their controls for the 3 studies. “Original study” is the original published study from Greater Manchester. “National analysis 1” is the first replication analysis using COVID-19 test data from the primary care data feed. “National analysis 2” is the second replication analysis using the Second-Generation Surveillance System for the COVID-19 test results. Values are presented as either “mean (standard deviation)” or “count (percentage)”.

|  | Original study |  | National analysis 1 |  | National analysis 2 |  |
| --- | --- | --- | --- | --- | --- | --- |
| Variable | Controls | T1D | Controls | T1D | Controls | T1D |
| **N** | 2573 (100%) | 862 (100%) | 114790 (100%) | 38523 (100%) | 223995 (100%) | 77392 (100%) |
| **Admission (within 28 days)** | 120 (5%) | 86 (10%) | 3735 (3%) | 3422 (9%) | 8665 (4%) | 8263 (11%) |
| **Age (years)** | 39.0 (17.0) | 39.4 (17.4) | 40.3 (18.0) | 40.5 (18.2) | 38.6 (18.3) | 38.9 (18.4) |
| **Sex (Is Male)** | 1349 (52.4%) | 454 (52.7%) | 58290 (50.8%) | 19655 (51.0%) | 117304 (52.4%) | 40722 (52.6%) |
| **Townsend score (higher is more deprived)** | 0.9 (3.7) | 0.9 (3.6) | -0.1 (3.6) | -0.2 (3.5) | 0.0 (3.6) | -0.1 (3.5) |
| **Townsend quintile (higher is more deprived)** |  |  |  |  |  |  |
| **1 (least deprived)** | 447 (17%) | 135 (16%) | 24210 (21%) | 7889 (21%) | 46237 (21%) | 15612 (20%) |
| **2** | 364 (14%) | 126 (15%) | 23891 (21%) | 8335 (22%) | 46173 (21%) | 16372 (21%) |
| **3** | 430 (17%) | 150 (17%) | 22092 (19%) | 7686 (20%) | 42955 (19%) | 15548 (20%) |
| **4** | 564 (22%) | 202 (23%) | 21294 (19%) | 7340 (19%) | 42809 (19%) | 15136 (20%) |
| **5 (most deprived)** | 768 (30%) | 249 (29%) | 23303 (20%) | 7273 (19%) | 45821 (21%) | 14724 (19%) |
| **Latest BMI Value** | 27.9 (6.2) | 27.2 (5.8) | 27.9 (6.7) | 27.4 (6.1) | 27.6 (6.8) | 27.0 (6.1) |
| **Latest LDL cholesterol Value** | 2.9 (0.9) | 2.5 (1.0) | 2.9 (0.9) | 2.4 (0.9) | 2.9 (0.9) | 2.4 (0.9) |
| **Latest HDL cholesterol Value** | 1.4 (0.4) | 1.4 (0.4) | 1.4 (0.4) | 1.5 (0.5) | 1.4 (0.4) | 1.5 (0.4) |
| **Latest eGFR Value** | 82.3 (13.8) | 80.5 (18.1) | 81.1 (13.7) | 80.3 (19.4) | 80.6 (14.2) | 79.7 (20.3) |
| **Latest HbA1c Value** | 34.5 (8.8) | 67.6 (22.7) | 36.5 (4.2) | 66.8 (18.8) | 36.4 (4.2) | 67.3 (19.2) |
| **Latest total cholesterol Value** | 5.0 (1.0) | 4.6 (1.1) | 5.0 (1.1) | 4.5 (1.1) | 5.0 (1.1) | 4.5 (1.1) |
| **Current smoking status** |  |  |  |  |  |  |
| **Non-smoker** | 1800 (70%) | 593 (69%) | 98551 (86%) | 32924 (86%) | 191091 (85%) | 65661 (85%) |
| **Smoker** | 773 (30%) | 269 (31%) | 16239 (14%) | 5599 (15%) | 32904 (15%) | 11731 (15%) |
| **Patient has asthma** | 430 (17%) | 149 (17%) | 19453 (17%) | 6583 (17%) | 35532 (16%) | 12782 (17%) |
| **Patient has COPD** | 41 (2%) | 21 (2%) | 1659 (1%) | 599 (2%) | 2940 (1%) | 1112 (1%) |
| **Patient has severe mental illness** | 41 (2%) | 21 (2%) | 921 (1%) | 387 (1%) | 1757 (1%) | 761 (1%) |
| **Patient has hypertension** | 257 (10%) | 197 (23%) | 11965 (10%) | 8580 (22%) | 20990 (9%) | 15869 (21%) |
| **Patient is on ACEI** | 178 (7%) | 210 (24%) | 5590 (5%) | 6805 (18%) | 9716 (4%) | 12537 (16%) |
| **Patient is on Aspirin** | 52 (2%) | 91 (11%) | 2417 (2%) | 3351 (9%) | 4098 (2%) | 6182 (8%) |
| **Patient is on Clopidogrel** | 27 (1%) | 37 (4%) | 1038 (1%) | 1215 (3%) | 1922 (1%) | 2361 (3%) |
| **Patient is on Metformin** | 9 (0%) | 117 (14%) | 224 (0%) | 4133 (11%) | 347 (0%) | 7418 (10%) |
| **Hospital length of stay (days)** | 3.8 (8.6) | 5.2 (8.2) | 2.8 (8.2) | 4.0 (9.2) | 2.6 (9.1) | 3.8 (10.3) |
| **Ethnicity** |  |  |  |  |  |  |
| **White** | 1731 (67%) | 650 (75%) | 95942 (84%) | 34755 (90%) | 185136 (83%) | 69364 (90%) |
| **Asian** | 334 (13%) | 73 (9%) | 8977 (8%) | 1701 (4%) | 17233 (8%) | 3373 (4%) |
| **Black** | 54 (2%) | 26 (3%) | 3113 (3%) | 869 (2%) | 6649 (3%) | 1950 (3%) |
| **Mixed** | 46 (2%) | 10 (1%) | 2128 (2%) | 610 (2%) | 4524 (2%) | 1346 (2%) |
| **Other** | 91 (4%) | 33 (4%) | 2497 (2%) | 455 (1%) | 5325 (2%) | 969 (1%) |
| **Unknown** | 317 (12%) | 70 (8%) | 2133 (2%) | 133 (0%) | 5128 (2%) | 390 (1%) |

**Table 2.** characteristics of the individuals with type 2 diabetes (T2D) and their controls for the 3 studies. “Original study” is the original published study from Greater Manchester. “National analysis 1” is the first replication analysis using COVID-19 test data from the primary care data feed. “National analysis 2” is the second replication analysis using the Second-Generation Surveillance System for the COVID-19 test results. Values are presented as either “mean (standard deviation)” or “count (percentage)”.

|  | **Original study** |  | **National analysis 1** |  | **National analysis 2** |  |
| --- | --- | --- | --- | --- | --- | --- |
| **Variable** | **Controls** | **T2D** | **Controls** | **T2D** | **Controls** | **T2D** |
| **N** | 37979 (100%) | 13225 (100%) | 1298984 (100%) | 448829 (100%) | 2354775 (100%) | 836532 (100%) |
| **Admission (within 28 days)** | 4407 (12%) | 2160 (16%) | 116443 (9%) | 68659 (15%) | 254496 (11%) | 155796 (19%) |
| **Age** | 62.2 (14.4) | 63.1 (14.4) | 62.8 (14.7) | 63.3 (14.7) | 63.0 (14.8) | 63.5 (14.9) |
| **Sex (Is Male)** | 20688 (54.5%) | 7427 (56.2%) | 675455 (52%) | 238400 (53%) | 1257080 (53.4%) | 454235 (54.3%) |
| **Townsend score (higher is more deprived)** | 0.4 (3.7) | 1.8 (3.7) | -0.6 (3.4) | 0.5 (3.7) | -0.6 (3.3) | 0.6 (3.7) |
| **Townsend quintile (higher is more deprived)** |  |  |  |  |  |  |
| **1** | 7540 (20%) | 1534 (12%) | 325211 (25%) | 75668 (17%) | 583601 (25%) | 137328 (16%) |
| **2** | 6126 (16%) | 1491 (11%) | 301249 (23%) | 83326 (19%) | 546987 (23%) | 153864 (18%) |
| **3** | 6888 (18%) | 2076 (16%) | 253188 (20%) | 84480 (19%) | 464107 (20%) | 158645 (19%) |
| **4** | 8062 (21%) | 2996 (23%) | 219340 (17%) | 91425 (20%) | 404138 (17%) | 174275 (21%) |
| **5** | 9363 (25%) | 5128 (39%) | 199996 (15%) | 113930 (25%) | 355942 (15%) | 212420 (25%) |
| **Latest BMI Value** | 28.6 (6.1) | 31.7 (6.9) | 28.1 (6.2) | 31.9 (7.2) | 28.0 (6.1) | 31.7 (7.2) |
| **Latest LDL cholesterol Value** | 2.8 (1.0) | 2.2 (0.9) | 2.7 (1.0) | 2.2 (0.9) | 2.7 (1.0) | 2.2 (1.0) |
| **Latest HDL cholesterol Value** | 1.4 (0.4) | 1.2 (0.3) | 1.5 (0.4) | 1.2 (0.3) | 1.5 (0.4) | 1.2 (0.3) |
| **Latest eGFR Value** | 75.9 (15.7) | 75.3 (18.7) | 74.0 (16.0) | 73.5 (19.2) | 73.2 (16.3) | 72.5 (19.8) |
| **Latest HbA1c Value** | 36.1 (9.1) | 56.6 (21.0) | 38.0 (4.1) | 58.1 (17.5) | 38.0 (4.2) | 58.3 (17.6) |
| **Latest total cholesterol Value** | 4.9 (1.1) | 4.3 (1.2) | 4.9 (1.1) | 4.3 (1.2) | 4.9 (1.1) | 4.3 (1.2) |
| **Current smoking status** |  |  |  |  |  |  |
| **Non-smoker** | 22519 (59%) | 7774 (59%) | 1137301 (88%) | 390957 (87%) | 2044839 (87%) | 722813 (86%) |
| **Smoker** | 15460 (41%) | 5451 (41%) | 161683 (12%) | 57872 (13%) | 309936 (13%) | 113719 (14%) |
| **Patient has asthma** | 5867 (15%) | 2401 (18%) | 199605 (15%) | 85642 (19%) | 345564 (15%) | 153313 (18%) |
| **Patient has COPD** | 2566 (7%) | 1011 (8%) | 67251 (5%) | 31576 (7%) | 123297 (5%) | 59235 (7%) |
| **Patient has severe mental illness** | 1300 (3%) | 603 (5%) | 15902 (1%) | 10232 (2%) | 29230 (1%) | 20144 (2%) |
| **Patient has hypertension** | 11337 (30%) | 7380 (56%) | 392765 (30%) | 252621 (56%) | 714311 (30%) | 472596 (57%) |
| **Patient is on ACEI** | 7695 (20%) | 6537 (49%) | 165378 (13%) | 149107 (33%) | 298067 (13%) | 275760 (33%) |
| **Patient is on Aspirin** | 3079 (8%) | 2559 (19%) | 90899 (7%) | 72149 (16%) | 165549 (7%) | 135184 (16%) |
| **Patient is on Clopidogrel** | 1607 (4%) | 1042 (8%) | 48941 (4%) | 31870 (7%) | 89827 (4%) | 60763 (7%) |
| **Patient is on Metformin** | 253 (1%) | 8150 (62%) | 1425 (0%) | 270421 (60%) | 2632 (0%) | 496184 (59%) |
| **Hospital length of stay (days)** | 6.7 (13.2) | 8.2 (16.2) | 5.3 (11.1) | 6.4 (12.0) | 5.1 (11.3) | 6.2 (12.4) |
| **Ethnicity** |  |  |  |  |  |  |
| **White** | 29405 (77%) | 7981 (60%) | 1157194 (89%) | 340211 (76%) | 2093541 (89%) | 632016 (76%) |
| **Asian** | 3082 (8%) | 3274 (25%) | 67877 (5%) | 73277 (16%) | 115543 (5%) | 133818 (16%) |
| **Black** | 833 (2%) | 498 (4%) | 27301 (2%) | 19576 (4%) | 51998 (2%) | 39550 (5%) |
| **Mixed** | 279 (1%) | 148 (1%) | 11411 (1%) | 5836 (1%) | 21204 (1%) | 11180 (1%) |
| **Other** | 1101 (3%) | 541 (4%) | 18073 (1%) | 7636 (2%) | 32759 (1%) | 14335 (2%) |
| **Unknown** | 3279 (9%) | 783 (6%) | 17128 (1%) | 2293 (1%) | 39730 (2%) | 5633 (1%) |

Most factors analysed were comparable with a few exceptions. Smoking status was much lower nationally (14-15% vs 30-31% for T1D, 12-14% vs 41% for T2D), but this was due to a categorisation error in the original study where anyone with a history of smoking was counted as a smoker. Greater Manchester is more ethnically diverse, but the GM data also has a higher proportion of unknown ethnicities, possibly because in the national SDE there are more sources of demographic data from which to determine an individuals’ ethnicity. Finally, patients in the national analyses had, on average, shorter lengths of stay in hospital. This is likely due to the later cut-off date for the national analyses, where the combined effect of the reduced severity of later strains, and the vaccination programme, mean that later diagnoses of COVID-19 are less likely to be severe.

### 3.2 T1D univariable analyses

Out of 25 variables analysed, only three (ACE inhibitor, metformin, or mixed ethnicity) showed a statistically significant difference in effect size between GM and the national data (Table S6). Mixed ethnicity had extremely small numbers in the GM study so the discrepancy here is likely due to random chance and the inconsistencies in reporting mixed ethnicity. For prescribed medications it is possible that not all metformin or ACE inhibitor SNOMED codes are extracted in the GDPPR dataset which may explain this discrepancy.

All variables that were statistically significant in the original study had the same positive or negative association (odds ratio direction) with the outcome in both national analyses (Figure 1).

**Figure 1.**
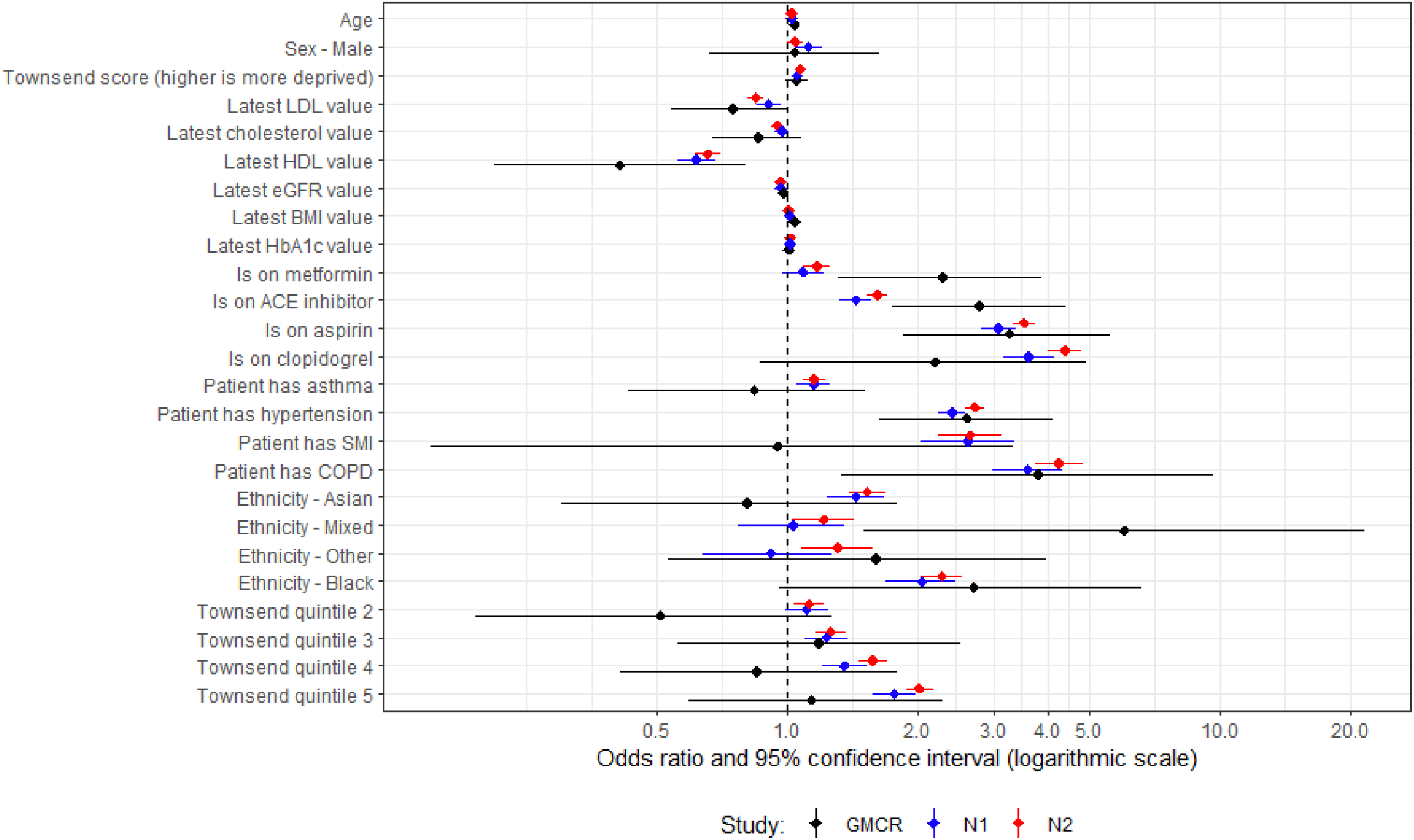
Univariable analysis for patients with type 1 diabetes. “GMCR” is the original published study (Greater Manchester Care Record), “N1” is the first replication analysis using COVID-19 test data from the primary care data feed, and “N2” is the second replication analysis utilising the Second-Generation Surveillance System for the COVID-19 test results.

### 3.3 T2D univariable analyses

For the first national analysis, out of 25 variables analysed, only four (latest HDL, COPD, ACE inhibitor, Townsend quintile 2) showed a statistically significant difference in effect size between GM and the national data (Table S6). For the second national analysis there were eight that showed a difference (age, cholesterol, eGFR, COPD, ACE inhibitor, clopidogrel, aspirin, Townsend quintile 2).

All results that were statistically significant in the original study had the same positive or negative association with the outcome in both national analyses (Figure 2).

**Figure 2.**
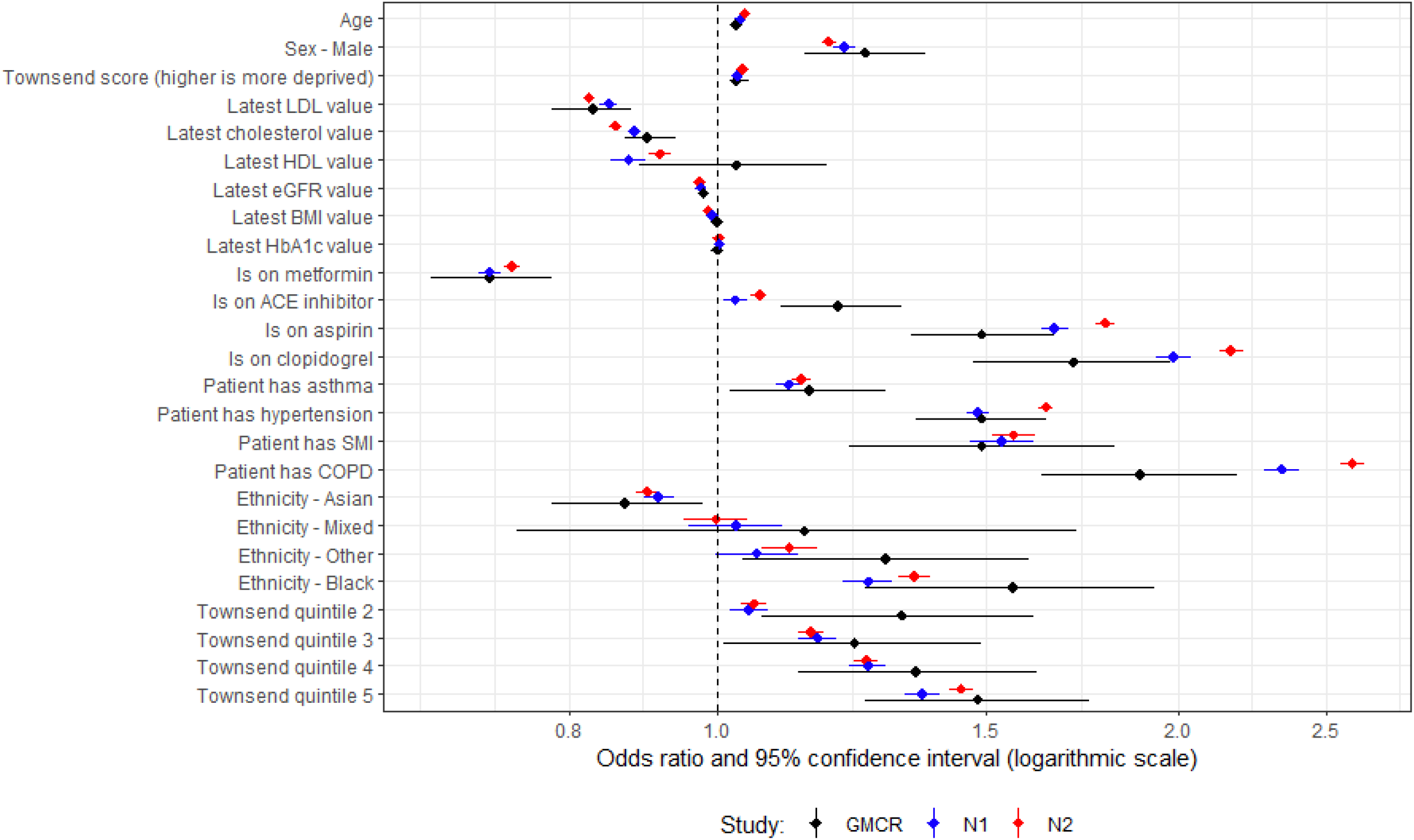
Univariable analysis for patients with type 2 diabetes. “GMCR” is the original published study (Greater Manchester Care Record), “N1” is the first replication analysis using COVID-19 test data from the primary care data feed, and “N2” is the second replication analysis utilising the Second-Generation Surveillance System for the COVID-19 test results.

### 3.4 T1D multivariable analyses

History of COPD, and mixed ethnicity were the only variables with a statistically significant difference in effect size between GM and the national data (Table S7). As mentioned earlier, the original study had very few patients coded as mixed ethnicity and so had a wide confidence interval, and while the ORs do not fall within the original CI, the CIs do overlap (Figure 3).

**Figure 3.**
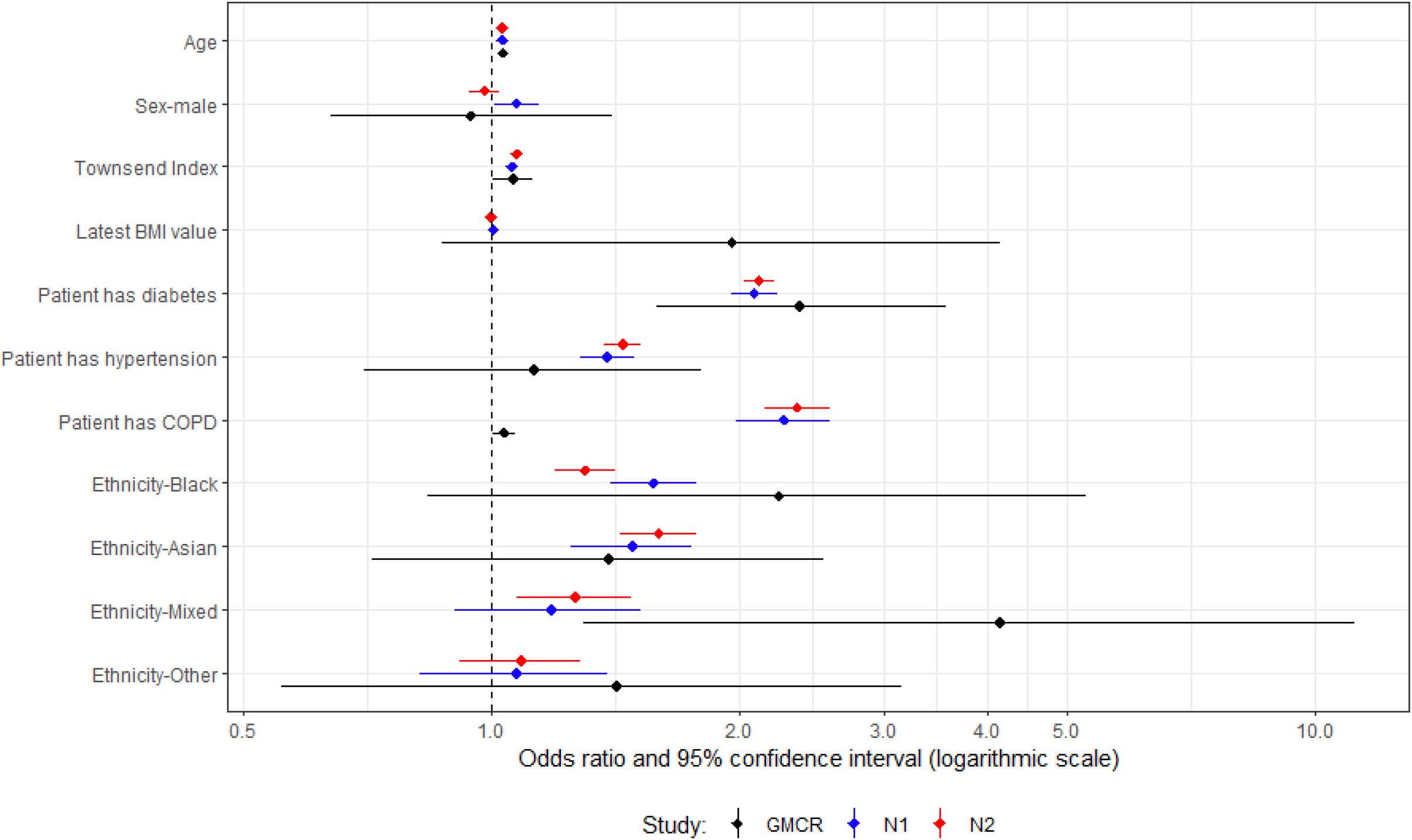
Multivariable analysis for patients with type 1 diabetes and their controls. “GMCR” is the original published study (Greater Manchester Care Record), “N1” is the first replication analysis using COVID-19 test data from the primary care data feed, and “N2” is the second replication analysis using the Second-Generation Surveillance System for the COVID-19 test results.

### 3.5 T2D multivariable analyses

For the first national analysis, eight (out of 11), and for the second, six (out of 11) variables showed a statistically significant difference in effect size between GM and the national data (Table S7).

Most variables have an OR in the national analyses that is outside the CI of the original study (Figure 4). However, all ORs are in the same direction as in the original study. Age, Townsend index, and hypertension all have a small, but significant, effect in all three analyses. Being male, or non-white ethnicity, have large effect sizes in all three analyses, though black ethnicity has a smaller odds ratio in the national analyses (first national OR=1.25 and second national OR=1.26 vs GM OR=1.79). Patients with diabetes and patients with COPD have a much larger OR in the national analyses (diabetes: 1.29 and 1.36 vs 1.1, COPD: 1.87 and 1.99 vs 1.03). Latest BMI has much smaller ORs in the national analyses (BMI: 1.03 and 1.02 vs 1.64).

**Figure 4.**
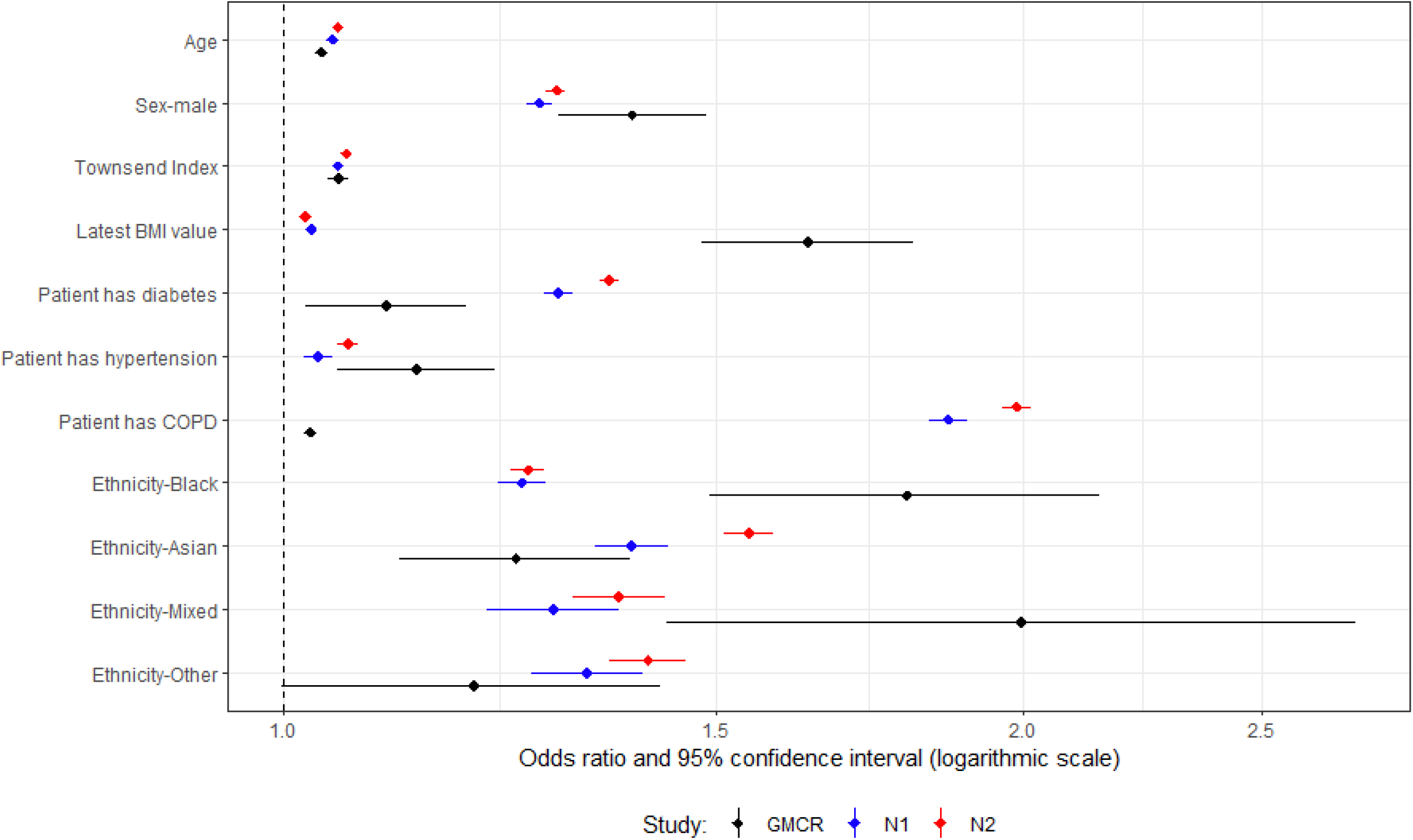
Multivariable analysis for patients with type 2 diabetes and their controls. “GMCR” is the original published study (Greater Manchester Care Record), “N1” is the first replication analysis using COVID-19 test data from the primary care data feed, and “N2” is the second replication analysis using the Second-Generation Surveillance System for the COVID-19 test results.

## 4 Discussion

We have conducted a study to determine the extent to which results replicate between a regional and a national database of electronic healthcare record data.

EHR data can be variable in quality and contain many unknowns and challenges [11]. However, they are typically analysed in large quantities which to some extent mitigates the effects of missingness and noise from random bias. Our analysis has shown that, while the actual odds ratios from multiple studies may vary, the direction and approximate magnitude of the effect size remains the same. All results that were significant remained significant, and therefore clinical decisions made on the results in the regional study would be consistent with the national analyses. This provides some evidence that the findings of regional studies can be extrapolated to a national setting.

However, there were also discrepancies, particularly in the multivariable analysis of patients with T2D and their controls. The large effect size of BMI in the original studies was much lower in the national analyses, and the effect of a patient having diabetes or COPD was much higher in the national analyses. The differences may be due to the underlying data sources, or to differences in the phenotypes as in the GM data the clinical coding was a mixture of Read v2, CTV3 and EMIS codes, while the national database was SNOMED. Therefore, it is important to replicate observational studies in different datasets to better understand the results due to genuine differences between the populations rather than those that are artefacts of the data.

Another explanation for discrepancies could be differences or mistakes in the data curation code. The data analysis code was the same in all studies, but the data curation code was different due to differences in the underlying data. All code used in this analysis is publicly available and therefore open to scrutiny, but it is hard to discover errors without in-depth examination. One option to discover such errors is for an independent study team to perform the same analysis on the same data. Reproduction of studies using the same data, but performed by a different study team would be beneficial. However, even that is not a panacea, as discovered in a recent study where 174 independent teams were given the same data and the same research question, and yet there was substantial heterogeneity among findings with some showing results with opposite associations with the outcome variable [12].

The cohort in the second national analysis was approximately double the cohort for the first national analysis for both T1D (77,392 patients vs 38,523) and T2D (836,532 vs 448,829). The difference between these cohorts was the addition of the SGSS dataset to identify more COVID-19 positive tests. SGSS is a much better source of COVID-19 test data, however there is no real difference between the results in the two national analyses suggesting that COVID-19 tests in the primary care record are sufficient for most research.

The original study population appears to have a higher proportion of severe mental illness (SMI) when compared with the national population. The prevalence in GM is likely to be higher than nationally. However, in this case it is predominantly because not all clinical codes used in the original analysis to define SMI were available in the GDPPR data set and so the apparent prevalence was lower nationally. The original study also had a much higher proportion of smokers. However, this was due to an error where patients who had ever had a current-smoker clinical code in their record were counted as smokers, even if they subsequently had quit. Smoking was therefore excluded from the replication study.

### 4.1 Strengths and limitations

The sources of data, while similar were different. GMCR has full GP data, while the GDPPR dataset in the national database is restricted to a subset of SNOMED codes. While reasonably comprehensive, it does not cover codes outside the scope of the GDPPR extract and there were three covariates in the original analysis that could not be used: vitamin D, testosterone and sex hormone binding globulin. Hospitalisation, the main outcome factor, was sourced from individual hospital feeds in the GMCR, while in the national database it was from the standardised HES data. However, despite these limitations the results were remarkably similar. It could also be argued that these are not limitations, and in fact demonstrate the challenges of replication even in two similar databases.

There are other limitations. Firstly, the findings in this replication study for this particular disorder may not be transferable to other conditions. However, it is likely to be similar for other long-term conditions diagnosed in primary care. Secondly, the same researchers conducted both studies and so may have made the same conceptual or procedural errors in both studies. In addition, knowing the previous study’s results may have subconsciously led us to confirm the previous findings rather than attempt to challenge them. However, in this case the replication benefitted from a mix of original researchers and new colleagues from the national SDE which ensured the replication was as objective as possible.

## 5 Conclusion

In two replication studies, performed in a national database, we have shown similar results with a previous study in a smaller, regional database. This provides evidence that results in regional databases can be extrapolated to national settings. However, there were still differences, which further highlights the need for replication of observational studies using electronic health record data, and for different study teams to reproduce work using the same data.

## 6 Author Statement

RW processed and cleaned the data, performed the analysis and drafted the manuscript. DJ designed and performed the analysis and reviewed the manuscript.

TB assisted with the data processing and cleaning and reviewed the manuscript.

AH provided clinical support, guidance on the analysis and discussion, and reviewed the manuscript. MM assisted with the data processing and cleaning and reviewed the manuscript.

MS designed and performed the analysis and reviewed the manuscript.

NP provided overall direction, guidance on the analysis and discussion, and reviewed the manuscript.

Members of the wider CVD-COVID-UK/COVID-IMPACT consortium (https://www.hdruk.ac.uk/wp-content/uploads/2021/12/211220-CVD-COVID-UK-COVID-IMPACT-Consortium-Members.pdf) also provided comments on drafts of the protocol and manuscript.

## 8 Ethical approval

The North East -Newcastle and North Tyneside 2 research ethics committee provided ethical approval for the CVD-COVID-UK/COVID-IMPACT research programme (REC No 20/NE/0161) to access, within secure trusted research environments, unconsented, whole-population, de-identified data from electronic health records collected as part of patients’ routine healthcare.

## 9 Acknowledgements

This work is carried out with the support of the BHF Data Science Centre led by HDR UK (BHF Grant no. SP/19/3/34678). This study makes use of de-identified data held in NHS England’s SDE for England, and made available via the BHF Data Science Centre’s CVD-COVID-UK/COVID-IMPACT consortium. This work uses data provided by patients and collected by the NHS as part of their care and support. We would also like to acknowledge all data providers who make health relevant data available for research.

## 11 Data availability

The data used in this study are available in NHS England’s SDE service for England, but as restrictions apply they are not publicly available (https://digital.nhs.uk/coronavirus/coronavirus-data-services-updates/trusted-research-environment-service-for-england). The CVD-COVID-UK/COVID-IMPACT programme led by the BHF Data Science Centre (https://bhfdatasciencecentre.org) received approval to access data in NHS England’s SDE service for England from the Independent Group Advising on the Release of Data (IGARD) (https://digital.nhs.uk/about-nhs-digital/corporate-information-and-documents/independent-group-advising-on-the-release-of-data) via an application made in the Data Access Request Service (DARS) Online system (ref. DARS-NIC-381078-Y9C5K) (https://digital.nhs.uk/services/data-access-request-service-dars/dars-products-and-services). The CVD-COVID-UK/COVID-IMPACT Approvals & Oversight Board (https://bhfdatasciencecentre.org/areas/cvd-covid-uk-covid-impact/) subsequently granted approval to this project to access the data within NHS England’s SDE service for England. The deidentified data used in this study were made available to accredited researchers only. Those wishing to gain access to the data should contact in the first instance.

## References

[1] M.D. Rotelli, Ethical Considerations for Increased Transparency and Reproducibility in the Retrospective Analysis of Health Care Data, Ther. Innov. Regul. Sci. 49 (2015) 342–347. 10.1177/2168479015578155.

[2] L.G. Hemkens, D.G. Contopoulos-Ioannidis, J.P.A. Ioannidis, Agreement of treatment effects for mortality from routinely collected data and subsequent randomized trials: Meta-epidemiological survey, BMJ. 352 (2016). 10.1136/bmj.i493.

[3] S.N. Goodman, D. Fanelli, J.P.A. Ioannidis, What does research reproducibility mean?, in: Get. to Good Res. Integr. Biomed. Sci., Springer International Publishing, 2018: pp. 96–102. 10.1126/scitranslmed.aaf5027.

[4] A.H. Heald, D.A. Jenkins, R. Williams, M. Sperrin, H. Fachim, R.N. Mudaliar, A. Syed, A. Naseem, J.M. Gibson, K.A. Bowden Davies, N. Peek, S.G. Anderson, Y. Peng, W. Ollier, The Risk Factors Potentially Influencing Hospital Admission in People with Diabetes, Following SARS-CoV-2 Infection: A Population-Level Analysis, Diabetes Ther. 13 (2022) 1007–1021. 10.1007/s13300-022-01230-2.

[5] R. Williams, T. Bolton, D. Jenkins, M.A. Mizani, M. Sperrin, C. Sudlow, A. Wood, A. Heald, N. Peek, The challenges of replication: a worked example of methods reproducibility using electronic health record data, MedRxiv. (2024) 2024.08.06.24311535. 10.1101/2024.08.06.24311535.

[6] NHS Digital, General Practice Extraction Service (GPES) Data for pandemic planning and research: a guide for analysts and users of the data, (n.d.). https://digital.nhs.uk/coronavirus/gpes-data-for-pandemic-planning-and-research/guide-for-analysts-and-users-of-the-data (accessed May 31, 2023).

[7] NHS Digital, Hospital Episode Statistics (HES), (n.d.). https://digital.nhs.uk/data-and-information/data-tools-and-services/data-services/hospital-episode-statistics (accessed June 29, 2023).

[8] NHS Digital, COVID-19 Second Generation Surveillance System (SGSS), (n.d.). https://digital.nhs.uk/services/data-access-request-service-dars/dars-products-and-services/data-set-catalogue/covid-19-second-generation-surveillance-system-sgss (accessed June 29, 2023).

[9] A. Wood, R. Denholm, S. Hollings, J. Cooper, S. Ip, V. Walker, S. Denaxas, A. Akbari, A. Banerjee, W. Whiteley, A. Lai, J. Sterne, C. Sudlow, Linked electronic health records for research on a nationwide cohort of more than 54 million people in England: Data resource, BMJ. 373 (2021). 10.1136/bmj.n826.

[10] A.H. Heald, D.A. Jenkins, R. Williams, R.N. Mudaliar, A. Khan, A. Syed, N. Sattar, K. Khunti, A. Naseem, K.A. Bowden-Davies, J.M. Gibson, W. Ollier, Sars-Cov-2 Infection in People with Type 1 Diabetes and Hospital Admission: An Analysis of Risk Factors for England, Diabetes Ther. 14 (2023) 2031–2042. 10.1007/s13300-023-01456-8.

[11] S. de Lusignan, N. Hague, J. van Vlymen, P. Kumarapeli, Routinely-collected general practice data are complex, but with systematic processing can be used for quality improvement and research., Inform. Prim. Care. 14 (2006) 59–66.

[12] E. Gould, H.S. Fraser, T.H. Parker, S. Nakagawa, S.C. Griffith, P.A. Vesk, F. Fidler, D.G. Hamilton, R.N. Abbey-Lee, J.K. Abbott, L.A. Aguirre, C. Alcaraz, I. Aloni, D. Altschul, K. Arekar, J.W. Atkins, J. Atkinson, C. Baker, M. Barrett, K. Bell, S.K. Bello, I. Beltrán, B.J. Berauer, M.G. Bertram, P.D. Billman, C.K. Blake, S. Blake, L. Bliard, A. Bonisoli-Alquati, T. Bonnet, C.N.M. Bordes, A.P.H. Bose, T. Botterill-James, M.A. Boyd, S.A. Boyle, T. Bradfer-Lawrence, J. Bradham, J.A. Brand, M.I. Brengdahl, M. Bulla, L. Bussière, E. Camerlenghi, S.E. Campbell, L.L.F. Campos, A. Caravaggi, P. Cardoso, C.J.W. Carroll, T.A. Catanach, X. Chen, H.Y.J. Chik, E.S. Choy, A.P. Christie, A. Chuang, A.J. Chunco, B.L. Clark, A. Contina, G.A. Covernton, M.P. Cox, K.A. Cressman, M. Crotti, C.D. Crouch, P.B. D’Amelio, A.A. de Sousa, T.F. Döbert, R. Dobler, A.J. Dobson, T.S. Doherty, S.M. Drobniak, A.G. Duffy, A.B. Duncan, R.P. Dunn, J. Dunning, T. Dutta, L. Eberhart-Hertel, J.A. Elmore, M.M. Elsherif, H.M. English, D.C. Ensminger, U.R. Ernst, S.M. Ferguson, E. Fernández-Juricic, T. Ferreira-Arruda, J. Fieberg, E.A. Finch, E.A. Fiorenza, D.N. Fisher, A. Fontaine, W. Forstmeier, Y. Fourcade, G.S. Frank, C.A. Freund, E. Fuentes-Lillo, S.L. Gandy, D.G. Gannon, A.I. García-Cervigón, A.C. Garretson, X. Ge, W.L. Geary, C. Géron, M. Gilles, A. Girndt, D. Gliksman, H.B. Goldspiel, D.G.E. Gomes, M.K. Good, S.C. Goslee, J.S. Gosnell, E.M. Grames, P. Gratton, N.M. Grebe, S.M. Greenler, M. Griffioen, D.M. Griffith, F.J. Griffith, J.J. Grossman, A. Güncan, S. Haesen, J.G. Hagan, H.A. Hager, J.P. Harris, N.D. Harrison, S.S. Hasnain, J.C. Havird, A.J. Heaton, M.L. Herrera-Chaustre, T.J. Howard, B.-Y. Hsu, F. Iannarilli, E.C. Iranzo, E.N.K. Iverson, S.O. Jimoh, D.H. Johnson, M. Johnsson, J. Jorna, T. Jucker, M. Jung, I. Kacergyte, O. Kaltz, A. Ke, C.D. Kelly, K. Keogan, F.W. Keppeler, A.K. Killion, D. Kim, D.P. Kochan, P. Korsten, S. Kothari, J. Kuppler, J.M. Kusch, M. Lagisz, K.M. Lalla, D.J. Larkin, C.L. Larson, K.S. Lauck, M.E. Lauterbur, A. Law, D.-J. Léandri-Breton, J.J. Lembrechts, K. L’Herpiniere, E.J.P. Lievens, D.O. de Lima, S. Lindsay, M. Luquet, R. MacLeod, K.H. Macphie, K. Magellan, M.M. Mair, L.E. Malm, S. Mammola, C.P. Mandeville, M. Manhart, L.M. Manrique-Garzon, E. Mäntylä, P. Marchand, B.M. Marshall, C.A. Martin, D.A. Martin, J.M. Martin, A.R. Martinig, E.S. McCallum, M. McCauley, S.M. McNew, S.J. Meiners, T. Merkling, M. Michelangeli, M. Moiron, B. Moreira, J. Mortensen, B. Mos, T.O. Muraina, P.W. Murphy, L. Nelli, P. Niemelä, J. Nightingale, G. Nilsonne, S. Nolazco, S.S. Nooten, J.L. Novotny, A.B. Olin, C.L. Organ, K.L. Ostevik, F.X. Palacio, M. Paquet, D.J. Parker, D.J. Pascall, V.J. Pasquarella, J.H. Paterson, A. Payo-Payo, K.M. Pedersen, G. Perez, K.I. Perry, P. Pottier, M.J. Proulx, R. Proulx, J.L. Pruett, V. Ramananjato, F.T. Randimbiarison, O.H. Razafindratsima, D.J. Rennison, F. Riva, S. Riyahi, M.J. Roast, F.P. Rocha, D.G. Roche, C. Román-Palacios, M.S. Rosenberg, J. Ross, F.E. Rowland, D. Rugemalila, A.L. Russell, S. Ruuskanen, P. Saccone, A. Sadeh, S.M. Salazar, K. Sales, P. Salmón, A. Sánchez-Tójar, L.P. Santos, F. Santostefano, H.T. Schilling, M. Schmidt, T. Schmoll, A.C. Schneider, A.E. Schrock, J. Schroeder, N. Schtickzelle, N.L. Schultz, D.A. Scott, M.P. Scroggie, J.T. Shapiro, N. Sharma, C.L. Shearer, D. Simón, M.I. Sitvarin, F.L. Skupien, H.L. Slinn, G.P. Smith, J.A. Smith, R. Sollmann, K.S. Whitney, S.M. Still, E.F. Stuber, G.F. Sutton, B. Swallow, C.C. Taff, E. Takola, A.J. Tanentzap, R. Tarjuelo, R.J. Telford, C.J. Thawley, H. Thierry, J. Thomson, S. Tidau, E.M. Tompkins, C.M. Tortorelli, A. Trlica, B.R. Turnell, L. Urban, S. Van de Vondel, J.E.M. van der Wal, J. Van Eeckhoven, F. van Oordt, K.M. Vanderwel, M.C. Vanderwel, K.J. Vanderwolf, J. Vélez, D.C. Vergara-Florez, B.C. Verrelli, M.V. Vieira, N. Villamil, V. Vitali, J. Vollering, J. Walker, X.J. Walker, J.A. Walter, P. Waryszak, R.J. Weaver, R.E.M. Wedegärtner, D.L. Weller, S. Whelan, R.L. White, D.W. Wolfson, A. Wood, S.W. Yanco, J.D.L. Yen, C. Youngflesh, G. Zilio, C. Zimmer, G.M. Zimmerman, R.A. Zitomer, Same data, different analysts: variation in effect sizes due to analytical decisions in ecology and evolutionary biology, (2024). https://ecoevorxiv.org/repository/view/6000/ (accessed May 3, 2024).

